# Usability of the Mayo Test Drive remote self-administered web-based cognitive screening battery in adults ages 35 to 100 with and without cognitive impairment

**DOI:** 10.1101/2024.05.24.24307909

**Authors:** Jay S. Patel, Teresa J. Christianson, Logan T. Monahan, Ryan D. Frank, Winnie Z. Fan, John L. Stricker, Walter K. Kremers, Aimee J. Karstens, Mary M. Machulda, Julie A. Fields, Jason Hassenstab, Clifford R. Jack, Hugo Botha, Jonathan Graff-Radford, Ronald C. Petersen, Nikki H. Stricker

**Affiliations:** Division of Neurocognitive Disorders, Department of Psychiatry and Psychology, Mayo Clinic, Rochester, Minnesota, USA; Department of Information Technology, Mayo Clinic, Rochester, Minnesota, USA; Division of Biomedical Statistics and Informatics, Department of Quantitative Health Sciences, Mayo Clinic, Rochester, Minnesota, USA; Department of Neurology and Psychological & Brain Sciences, Washington University in St. Louis, St. Louis, Missouri, USA; Department of Radiology, Mayo Clinic, Rochester, Minnesota, USA; Department of Neurology, Mayo Clinic, Rochester, Minnesota, USA

**Keywords:** digital health, neuropsychology, smartphone, aging, telemedicine, computerized neuropsychological assessment, unsupervised, mild cognitive impairment, feasibility, adherence

## Abstract

**Background:** Self-administered cognitive screening measures that can be completed unsupervised and remotely have significant promise for high scalability. Mayo Test Drive (MTD): Test Development through Rapid Iteration, Validation and Expansion, is a web-based remote cognitive assessment platform for self-administered neuropsychological measures with previously demonstrated validity and reliability. Our primary study aim was to examine the usability of MTD. We additionally described participation and adherence rates; characterized factors that inform feasibility to implement MTD in research and clinical settings such as session and subtest durations, frequency of interruptions and noise during test sessions, device type use across demographic and clinical characteristics for completed sessions; and reported qualitative themes of free-text participant comments.

**Methods:** 1,950 Mayo Clinic Study of Aging and Mayo Alzheimer’s Disease Research Center participants (97% White, 99% Non-Hispanic) were invited to participate in this ancillary, uncompensated remote study. Most invitees were cognitively unimpaired (CU; *n*=1,769; 90.7%) and 9.3% were cognitively impaired (CI; *n*=181). Participation and adherence rates were defined as the number of participants initiating a test session when invited or due to complete a session, respectively. Usability was objectively defined as the percentage of participants who completed a session after initiating a session for a given timepoint (i.e., completion rates). MTD sessions included the Stricker Learning Span (SLS) learning and delay trials, four trials of the Symbols test, and completion of self-report questions that informed session context. All participants were invited to provide free-text comments that were coded by investigators and descriptively characterized by qualitative analysis.

**Results:** Baseline session completion rates were 98.5% across participants who initiated a test session (n=1199/1217, mean age 71, SD=12, range 35-100) and were comparable between CU (98.7%) and CI (95.0%) groups (*p*=.23). Completion rates did not significantly differ by age groups among CU participants (*p*>.10) and completion rates remained high in individuals 80+ (n=251, 97.3%). Participation rates were higher in the CU (n=1142, 65.4%) versus CI (n=57, 33.1%) group (*p*<.001). Participation rates were lower in those 80+ (53.8%) relative to younger age groups (68.9% ages 34-64; 70.2% ages 65-69). Adherence (i.e., retention) rate for a 7.5-month follow-up session was 89%. Average session duration was 16 minutes. Most participants used a personal computer (n=751, 62.7%), followed by a smartphone (n=266, 22.2%) or tablet (n=177, 14.8%). Comments were entered by 36.4% of participants and reflected several themes relevant to acceptability, face validity, usability, as well as comments informative for session context.

**Conclusions:** MTD demonstrated high usability as defined by completion rates in this research sample that includes a broad age range, though participation rates are lower in individuals with cognitive impairment. Results support good adherence at follow-up, feasibility through mean session durations, and acceptability based on qualitative analysis of participant comments.

## Introduction

Self-administered digital cognitive measures have the potential to increase access to cognitive testing relative to traditional person-administered cognitive screening and assessment [1]. The degree to which digital cognitive tests are usable, feasible and acceptable is important for successful implementation and scalability. For example, many digital tools are designed for a broad target population but ultimately require supervision and have high technological demands that result in a similar degree of administration resources as paper-and-pencil tests.

Amirpour et al. [2] tailored definitions of usability, feasibility, and acceptability for digital cognitive assessment technologies that we adopt for this study. Usability addresses the extent to which self-administered cognitive measures can be used by the target population and was characterized in this study through completion rates and participant feedback. Feasibility explores factors that may influence the successful implementation of the tool for the intended purpose in the target population, including user experience factors like duration, test session context, and device use. Acceptability reflects factors that may influence willingness of the target population to use a digital, self-administered cognitive assessment (e.g., participation rates, adherence/retention rates, and user feedback). Characterizing these factors for both person-administered and self-administered computerized measures is critical for identifying potential implementation and scalability barriers. For example, Hackett et al. [3] reported that the Picture Sequence Memory and List Sorting Working Memory subtests from the NIH Cognition Toolbox were too challenging for participants with cognitive impairment (i.e., low completion rates) and were ultimately removed from their study that used in-person, interactive cognitive testing. They also reported 21% of participants had missing data on remaining subtests from the NIH Cognition Toolbox. Thus, the utility of digital tools, regardless of a paradigm’s reliability and validity, can be limited by whether the target population is able and willing to engage with the testing platform and complete the test(s) as intended.

During the COVID-19 pandemic, Jacobs et al. [4] surveyed participants enrolled in an Alzheimer’s Disease Research Center and found that 84% of cognitively unimpaired (CU) individuals and 74% of individuals with cognitive impairment (but without dementia) were willing to engage in interactive, video-based remote cognitive assessment (e.g., telehealth). Most (85%) reported device access. In contrast only 39% of individuals with dementia reported willingness to engage in a single remote cognitive assessment and 52% reported device access. Slightly fewer participants expressed willingness to engage in multiple sessions of very brief cognitive testing (i.e., “burst” model) on a smartphone (66% CU, 59% cognitive impairment, 39% dementia). These survey results provide insights about attitudes toward remote cognitive testing agnostic of any specific platform but are limited to participant response rather than observed behavioral outcomes. Several studies have directly explored usability, feasibility, and acceptability in older-adult samples, with most demonstrating satisfactory usability rates [5–10]. However, adherence rates, defined as continued participation over longitudinal follow-up, decrease over time, limiting the utility of these remote assessments for longitudinal monitoring [8, 10–12]. Cognitive status, demographic variables, and subjective memory concerns influence elements of usability and acceptability including engagement, adherence, and retention rates [9, 12, 13].

Mayo Test Drive (MTD): Test Development through Rapid Iteration, Validation and Expansion was intentionally designed for high usability and ease of access. Design needs of older adults and individuals with cognitive impairment prioritized in platform development included low technology demands (e.g., no app download or log-in requirements, no swiping/dragging responses that are associated with decreased acceptability and usability [3, 14], no app navigation, no speaker/microphone use), multi-device compatibility, easy-to-follow instructions, visual optimization (e.g., high contrast displays, large font size), and consideration of the high base rate of hearing impairment in older adults [15]. MTD is a web-based platform that is compatible with a variety of devices, including smartphones, tablets, and personal computers. Users receive a unique, one-click url link with an embedded study name and participant ID. MTD has a simple user interface that only requires single touch (or click) responses. The platform can be used without personally identifiable information to decrease privacy concerns.

The MTD screening battery includes (1) a computer-adaptive word-list memory test, the Stricker Learning Span (SLS) [16], that shows similar ability to differentiate Alzheimer’s disease biomarker-defined groups as the in-person administered Rey’s Auditory Verbal Learning Test [17] and (2) a measure of processing speed/executive functioning that also requires visual discrimination (Symbols) [18, 19]. The MTD composite shows robust associations with an in-person administered Mayo Preclinical Alzheimer’s disease Cognitive Composite (Mayo-PACC)[20] and with core Alzheimer’s disease biomarkers, as well as large effect sizes for differentiating individuals with and without cognitive impairment [18]. Preliminary support for the feasibility, validity, and reliability of MTD was previously reported in an initial all-remote pilot study in 96 women aged 55-79 without dementia; 98% of participants completed a test session when a session was initiated [19].

The primary aim of the current study was to examine the usability of MTD in a large sample of adults and older adults. Usability was objectively defined as the percentage of participants who completed a session after initiating a session for a given timepoint (i.e., completion rates). We hypothesized that completion rates would be greater than 90%. We also explored whether completion rates differed by cognitive status and age. We included additional descriptive aims. First, we report participation rates to describe the demographic and clinical characteristics of individuals willing vs. not willing to initiate participation in an ancillary, uncompensated, online remote cognitive assessment study. Second, we report adherence (i.e., retention) rates for those due for a follow-up session. Third, we characterize factors that inform feasibility to implement MTD in research and clinical settings such as session and subtest durations (i.e., efficiency), frequency of interruptions and noise during test sessions, and device type use across demographic and clinical characteristics for completed sessions. Finally, we qualitatively analyzed themes relating to acceptability, usability and feasibility through voluntarily free-text user comments provided at the end of completed test sessions.

## Methods

### Participants and Recruitment Procedures

The Mayo Clinic Study of Aging (MCSA) is the primary source of participants for this study. The MCSA is a population-based study of individuals aged 30 years and older living in Olmsted County, MN who are randomly sampled to meet sex- and age-stratification goals using the resources of the Rochester Epidemiology Project medical records-linkage system [21]. Exclusion criteria are a terminal illness or hospice. Over 60% of residents contacted enroll in the MCSA and follow-up retention is 80%. Study visits include neurological examination with medical history review and administration of the Short Test of Mental Status (STMS) [22], clinical interview and completion of the Clinical Dementia Rating® scale by a study coordinator [23], and in-person neuropsychological testing [24]. After each study visit, a diagnosis of CU, MCI [25], or dementia [26] is established after the examining physician, interviewing study coordinator, and neuropsychologist make independent diagnostic determinations and then reach consensus agreement [24]. Prior visit data and MTD data are not considered for diagnosis. CU individuals aged 50 or older and participants with MCI or dementia complete in-person study visits every 15 months. CU individuals aged 50 or younger complete in-person study visits every 60 months. Additional participants were recruited from the Mayo Clinic Alzheimer’s Disease Research Center (ADRC; Rochester, MN).

Parent studies were approved by the Mayo Clinic Institutional Review Board (IRB) and the MCSA was additionally approved by the Olmsted Medical Center IRB. Written consent was obtained for participation in the parent study protocols (MCSA or ADRC) and oral consent was obtained for participation in the ancillary MTD remote study protocol (approved by Mayo Clinic). This study was conducted in accordance with the Declaration of Helsinki.

The MTD study in the MCSA and ADRC began with a pilot phase (5/25/21–9/3/21). A limited number of participants were initially invited to ensure study tasks could be completed by select groups that included newly enrolled (first visit) MCSA participants, individuals 70+ previously participating in a Cogstate Brief Battery (CBB) home-based option (for details, see [27], individuals aged 80+ not previously participating in a CBB home-based option, and individuals with cognitive impairment. During this phase, the study coordinator attempted to make phone calls to all participants who did not complete MTD after reminder emails. From 9/4/21-10/3/21 we invited all new MCSA enrollees, but no phone follow-up was provided. An initial phase of large-scale recruitment occurred 10/4/21-5/9/22, wherein all participants with MCSA visits (new and return) were invited to participate; limited phone follow-up and support was provided. As of 5/10/22 study coordinator support was increased so that we could continue to offer some phone follow-up support and a small number of in-clinic visits (upon request).

The primary recruitment method in the MCSA was via email. Participants were provided an MTD information sheet at the time of the in-person study visit that provided the study name and study contact information and alerted them that they may receive an email or phone call inviting them to participate in this study. An email invitation that explained the study and contained oral consent elements was sent the following week. Each participant received two reminder emails about one week apart, and participants were placed on a “to call” list if they did not respond to the final reminder email (when study coordinator resources allowed for phone call reminders). For any individuals requiring a legally authorized representative (LAR), an interactive consent conversation with the LAR and participant was completed.

ADRC recruitment started 6/10/21 and focused on individuals with MCI and dementia due to AD to increase representation of cognitively impaired participants because the MCSA includes predominantly CU participants. Because of this recruitment focus in the ADRC, the primary recruitment method for that parent study was an oral consent conversation at the time of the in-person visit. If consented, then emails with instructions and the test link were sent the following week. Of note, relatively few participants were recruited from the ADRC parent study due to prioritization of another NIH-funded study targeting similar participants during this time frame [28].

### MTD Procedures

MTD emails contained links that provided direct access to the assessment without any log-in requirements. Emails also included a QR code link; if clicked, users are taken to a website with a QR code that can be used if the participant is reading their email on a personal computer and would prefer to take the test on a mobile device (smartphone or tablet). Emails provided general instructions, including that participants should take the tests when they have 15-20 minutes in a quiet environment; that they can use a smartphone, tablet, or personal computer to take the tests; to complete the tests in one sitting; to avoid closing their web browser; and that they can receive help getting to the testing website or with any technological questions but that they should take the tests alone without any interruptions (e.g., no television, radio, or conversation). They are told not to share the email or the links with others or to allow someone else to take the test using their link. They are informed they will be invited to provide comments when they reach the end of testing.

Participants received additional instructions within the MTD test session after a brief welcome screen. Participants are again instructed to complete the tests by themselves in a quiet area where they will not be distracted for the 15-20 minutes and are again asked to complete the tests without direct assistance from another person. Participants report their location on the next screen (multiple-choice format). The test is considered “initiated” when participants make a response on this location-selection screen. For the current study, participants are considered to have completed the MTD session for a given timepoint if, after initiating a session, a session is subsequently fully completed.

The MTD screening battery consists of two subtests: the Stricker Learning Span (SLS) and the Symbols test. The SLS is a computer adaptive list learning task, previously described in detail [17–19, 29]. Briefly, single words are visually presented sequentially across five learning trials (range of words presented is 8-23 following adaptive rules) [17]. After each list presentation, memory for each word presented is tested with four-choice recognition. The delay trial of the SLS occurs following completion of the Symbols Test. The Symbols Test is an open-source measure of processing speed/executive functioning with previously demonstrated validity and reliability [18, 30, 31]. For each item, participants identify which of two symbol pairs on the bottom of the screen matches one of three symbol pairs presented at the top of the screen. There are four 12-item trials administered sequentially in MTD.

Both the SLS and Symbols start with a practice (called a “warm-up”). A single item is presented (single word to remember followed by 4-choice recognition or single Symbols item). If correct the first time, participants continue to the full subtest and are assigned a warm-up score of 4/4. If incorrect, another practice item is presented (score of 3/4 if passed on second attempt or 2/4 if passed on third attempt). If a participant cannot pass the warm-up within three attempts, the test is discontinued and there is one additional task to determine ability to follow basic instructions (1/4 if correct; 0/4 if incorrect).

Each subtest is followed by a question asking if anything interfered with performance during that test (SLS 1-5, Symbols, SLS Delay). If endorsed, a follow-up item presents several response options to select from. After all subtests are completed, participants are asked to self-report their device type, method of response input and whether it was noisy when they completed the tests. If noise is endorsed, a follow-up item presents several response options to select from.

Qualitative analyses of baseline sessions examined voluntary free-text data from comment boxes embedded in MTD at the end of completed sessions. All comments were first reviewed (NHS, JSP, LTM) and potential coding categories with example responses were generated and subsequently used to assign ratings. Comments were coded into the following categories: acceptability, face validity (a subset of acceptability), usability, and behavioral observations proxy. After initial training and double rating to ensure fidelity, LTM served as the primary rater. JSP and NHS reviewed ratings for accuracy. For comments that were hard to categorize or for which there was disagreement, consensus meetings were held to review rating options and reach group consensus (LTM, JSP, NHS, AJK).

### Inclusion Criteria

This study included MCSA and ADRC participants invited to complete MTD between 5/5/21 and 10/4/22. Available linked parent study data as of 11/13/22 were included, allowing a 6-week window to complete MTD following the first invitation. Nearly all participants completed MTD remotely except for 9 individuals who requested to come to clinic to complete MTD in-person for their baseline visit and 1 who requested to come to clinic for the follow-up (2^nd^) MTD session. Individuals completing MTD in clinic were assisted in initiating the test session and then were left in a quiet room alone to complete the test session but could request help if needed. All other participants self-administered MTD remotely. Participants were able to call or email a study coordinator for questions or assistance, as needed.

### Statistical Analyses

Data were descriptively summarized using means and standard deviations for continuous variables, and counts and percentages for categorical variables. Comparisons of data distributions across participation and completion status were performed using chi-square/Fisher exact tests for categorical variables (where appropriate), and linear regression models for continuous variables. *P* values adjusted for the effects of age, sex, and education were calculated from logistic regression models for dichotomous outcomes (clinical diagnosis), and multinomial logistic regression for categorical variables with 3 levels (categorized age). When categorized age was the outcome, we only adjusted for sex and education. All *P* values are 2-sided; all statistics were performed using SAS version 9.4 (SAS Institute Inc., Cary, NC).

## Results

### Participant characteristics

A total of 1,950 individuals were invited to participate in this study. Most invitees were from the Mayo Clinic Study of Aging (MCSA; 99%), and a subsample were from the Mayo Clinic Alzheimer’s Disease Research Center (ADRC; 1%). Mean age of invitees was 73 years, 50% were female, average education was 15 years, and invitees were predominately White (97%) and non-Hispanic/Latinx/e/o/a (99%; see **Table 1**). Most invitees were CU (*n*=1,769; 90.7%). Those diagnostically categorized as cognitively impaired (CI; *n*=181; 9.3%) consisted of participants with mild cognitive impairment (MCI; *n*=144) and dementia (*n*=36).

**Table 1.** Sociodemographics and cognitive status by group.

|  | Invitees<br>N=1950 | Non-Participants<br>N=733 | Participants<br>N=1217 | *P* | Non-Completers<br>N=18 | Completers<br>1199 | *P* |
| --- | --- | --- | --- | --- | --- | --- | --- |
| Sample size |  |  |  |  |  |  |  |
| **Age (years), Mean (SD)** | 72.7<br>(12.4) | 75.0 (13.5) | 71.3 (11.5) | <.001 | 80.1 (8.41) | 71.2 (11.5) | <.001 |
| Range | 34.4 -<br>101.1 | 34.4 – 101.1 | 35.8 –<br>100.3 |  | 62.76 – 89.7 | 35.8 –<br>100.3 |  |
| **Sex** |  |  |  | .462 |  |  | .147 |
| Female, n (%) | 968 (49.6) | 356 (48.6) | 612 (50.3) |  | 6 (33.3) | 606 (50.5) |  |
| Male, n (%) | 982 (50.4) | 377 (51.4) | 605 (49.7) |  | 12 (66.7%) | 593 (49.5) |  |
| **Education (years), Mean (SD)** | 15.3 (2.4) | 14.9 (2.5) | 15.6 (2.4) | <.001 | 15.6 (2.6) | 15.6 (2.4) | .931 |
| Range | 6 – 20 | 6.0 – 20.0 | 6 – 20 |  | 12-20 | 6 – 20 |  |
| **Race** |  |  |  | .396 |  |  | 1.00 |
| White, n (%) | 1883<br>(97.0) | 701 (96.3) | 1182 (97.4) |  | 18 (100.0) | 1164 (97.3) |  |
| Black, n (%) | 18 (0.9) | 10 (1.4) | 8 (0.7) |  | 0 (0) | 8 (0.7) |  |
| Asian, n (%) | 25 (1.3) | 12 (1.6) | 13 (1.1) |  | 0 (0) | 13 (1.1) |  |
| Native American/Alaskan, n (%) | 2 (0.1) | 1 (0.1) | 1 (0.1) |  | 0 (0) | 1 (0.1) |  |
| Native Hawaiian/Pacific Islander, n (%) | 1 (0.1) | 0 (0) | 1 (0.1) |  | 0 (0) | 1 (0.1) |  |
| More than one, n (%) | 13 (0.7) | 4 (0.5) | 9 (0.7) |  | 0 (0) | 9 (0.8) |  |
| Unknown/not rep, n (%) | 8 (0.4) | 5 (0.7) | 3 (0.2) |  | 0 (0) | 3 (0.3) |  |
| **Ethnicity** |  |  |  | .031 |  |  | 1.00 |
| Non-Hispanic/Latine, n (%) | 1935<br>(99.2) | 728 (99.3) | 1207 (99.2) |  | 18 (100.0) | 1189 (99.2) |  |
| Hispanic or Latine, n, (%) | 7 (0.4) | 0 (0) | 7 (0.6) |  | 0 (0) | 7 (0.6) |  |
| Unknown, n, (%) | 8 | 5 (0.7) | 3 (0.2) |  | 0 (0) | 3 (0.3) |  |
| **Kokmen Short Test of Mental Status, Mean (SD)** | 34.9 (3.3) | 34.0 (4.1) | 35.5 (2.6) | <.001 | 33.9 (4.6) | 35.5 (2.5) | .010 |
| Missing | 45 | 20 | 25 |  | 0 | 25 |  |
| Range | 1.0 – 38.0 | 1.0 – 38.0 | 12.0 – 38.0 |  | 19.0 – 38.0 | 12.0 – 38.0 |  |
| **Estimated Mini Mental Status Exam, Mean (SD) <sup>a</sup>** | 28.1 (2.1) | 27.6 (2.6) | 28.5 (1.5) | <.001 | 27.4 (2.9) | 28.5 (1.5) | .004 |
| Missing | 45 | 20 | 25 |  | 0 | 25 |  |
| Range | 1.0 – 30.0 | 1.0 – 30.0 | 12.0 – 30.0 |  | 18.0 – 30.0 | 12.0 – 30.0 |  |
| **CDR Global score** |  |  |  | <.001 |  |  | .242 |
| Missing | 11 | 6 | 5 |  | 1 | 4 |  |
| 0 (N, %) | 1766 (91.1) | 619 (85.1) | 1147 (94.6) |  | 15 (88.2) | 1132 (94.7) |  |
| 0.5 (N, %) | 154 (7.9) | 91 (12.5) | 63 (5.2) |  | 2 (11.8) | 61 (5.1) |  |
| 1 (N, %) | 11 (0.6) | 10 (1.4) | 1 (0.1) |  | 0 (0) | 1 (0.1) |  |
| 2 (N, %) | 8 (0.4) | 7 (1.0) | 1 (0.1) |  | 0 (0) | 1 (0.1) |  |

<sup>a</sup> MMSE score is estimated based on the Kokmen Short Test of Mental Status.
*Note:* Table used with permission of Mayo Foundation for Medical Education and Research; all rights reserved.

### Participation

Of all participants invited (*n*=1,950), 62.4% initiated an MTD session (*n*=1,217, i.e., participated). Most non-participants (67%) did not respond and 32.8 % declined (also see **Supplemental Table 1**). Participation rates were higher in the CU group (65.4%) compared to the cognitively impaired group (33.1%, *p* < 0.001). MTD participants (*n*=1217) were slightly younger (mean age 71.3 vs 75.0, *p* < .001) with slightly higher years of education (15.6 vs 14.9, *p* < .001) compared to those not participating (*n*=733; **Table 1**). Sex and race were not significantly different across participation groups (*p*’s > .40) though we note that all invited (*n*=7) Hispanic/Latinx/e/o/a participants did participate, resulting in a statistical difference across ethnicity groups (*p*=0.03). Participants scored slightly higher on the STMS global cognitive screening measure (35.5 vs 34.0, *p* < .001; estimated MMSE 28.5 vs 27.6, *p* < .001) [32] and were more likely to have CDR global score of zero compared to non-participants (94.6% vs 85.1%, *p* < 0.001).

Participation rates varied by age in the CU group (adjusted *p* <.001). Individuals between ages 34-64 and 65-79 demonstrated similar participation rates (68.9% and 70.2%, respectively), whereas the participation rate for those over 80 years was 53.8% **Supplemental Table 2**). Participation rates were not significantly different across MCI and dementia subgroups (34.9% vs 25.7%, adjusted *p* =0.30, **Supplemental Table 3**). Less than 1% of participants requested to come to the clinic to complete MTD (*n*=7); 99.4% of participants engaged in MTD remotely. **Usability**

Usability was examined through baseline completion rates. Most participants who initiated an MTD baseline test session completed a session (“completers”), with 98.5% overall completion rates. Completion rates were slightly higher in the CU (1142/1157=98.7%) than cognitively impaired (57/60=95.0%) groups, but this difference did not reach significance (adjusted *p* = 0.23; **Figure 1, Supplemental Table 1**). In comparison to non-completers (*n* = 18), completers (n = 1199) were younger (mean 71.2 vs 80.1, *p* = .001). There were no significant differences in other sociodemographic characteristics across completion groups (all *p*’s > 0.15). Completers scored slightly higher on a cognitive screening measure compared to non-completers (STMS: 35.5 vs 33.9, *p* = .01; estimated MMSE: 28.5 vs 27.4, *p* = .004). However, the CDR global score was not significantly different across completers and non-completers (*p* = .24).

**Figure 1.**
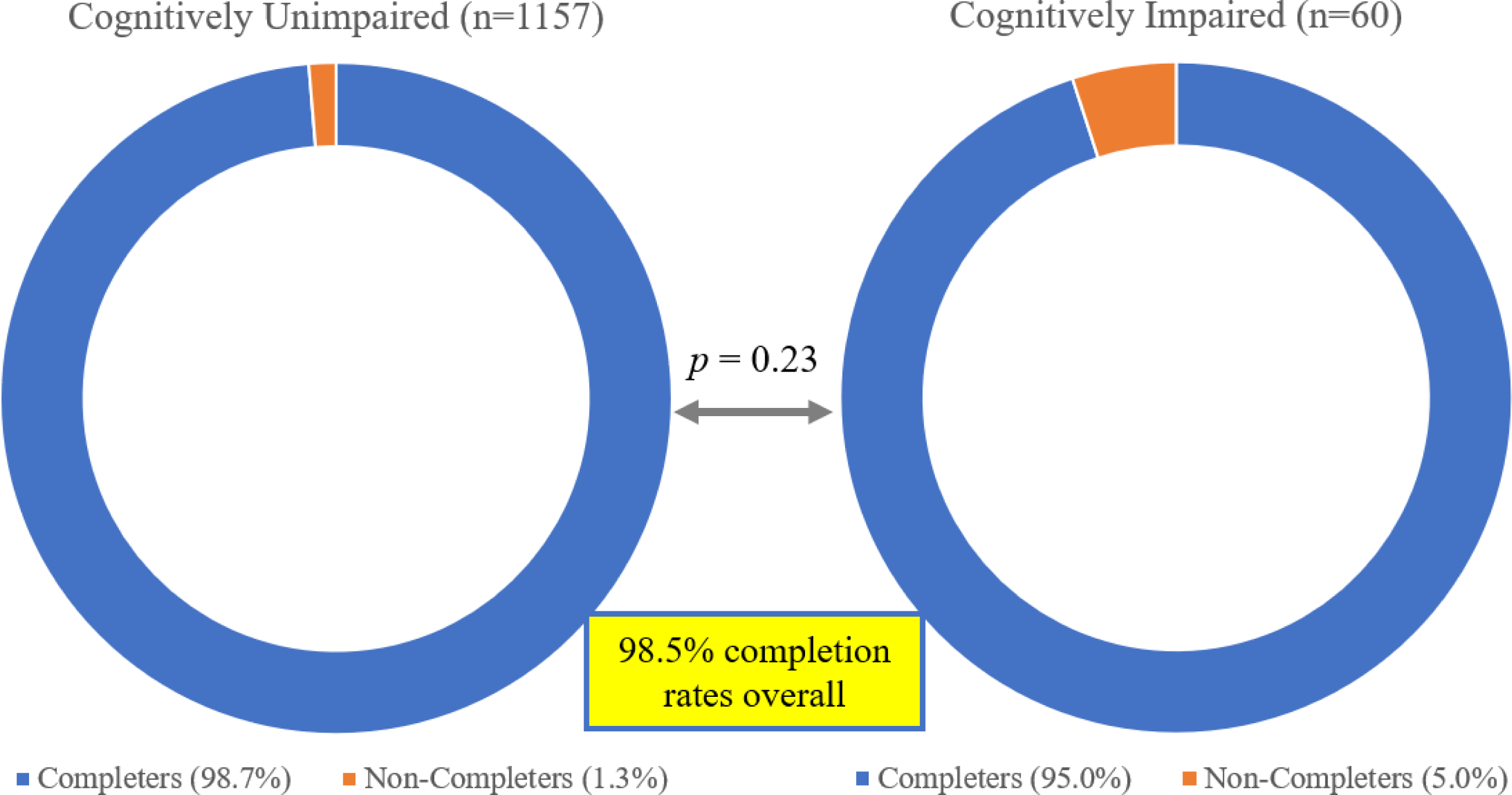
Usability: participant completion rates. *Note*. Figure used with permission of Mayo Foundation for Medical Education and Research, all rights reserved.

There were no significant differences in completion rates across age groups among CU participants (99.7%, 98.8%, and 97.3% in ages 34-64, 65-79, and 80+ respectively; adjusted *p*=0.10, **Supplemental Table 2**). Completion rates were not significantly different across MCI and dementia subgroups (96.1% vs 88.9%, *p* = .36, **Supplemental Table 3**).

### Adherence

A subsample of 583 participants were asked to complete a second session of MTD approximately 7-8 months after the baseline MTD session. Adherence (i.e., retention), defined as individuals who initiated a test session among those due for a follow-up session, was 89.0% (519/583) at follow-up. Completion rates at the follow-up session were high (99.0%, 514/519 of those initiating a follow-up session completed a session). See **Supplemental Table 4** for additional details.

### Feasibility

Mean session duration was 16.4 minutes (all completers, baseline session). The CU group demonstrated faster completion times (mean=16.2 minutes) than the CI group (mean=19.3 minutes, unadjusted *p* < .001), but this was likely due to the older age of the CI group, as this comparison was no longer significant after age/sex/education adjustment (adjusted *p* = 0.77).

SLS completion times were comparable across CU and CI groups (SLS Trials 1-5 mean: 8.7 minutes vs 8.7 minutes, adjusted *p* =0.74; SLS Delay mean: 1.4 minutes vs 1.5 minutes, adjusted *p* =0.68). Symbols completion was slower in the cognitively impaired group (mean=5.3 minutes) relative to the CU group (mean=3.5 minutes, adjusted *p* < .001; see **Table 2**). Among CU participants, older age was associated with longer duration times even after adjustment for sex and education (adjusted *p* < .001 for all; see **Table 3**).

**Table 2.** Session characteristics for all participants who completed a baseline MTD session by diagnostic subgroup.

|  | Total Completers<br>N=1199 | Cognitively<br>Unimpaired<br>N=1142 | Cognitively<br>Impaired<br>N=57 | Unadjusted<br>*P*<sup>a</sup> | Adjusted<br>*P*<sup>b</sup> |
| --- | --- | --- | --- | --- | --- |
| **Age at MTD (years), Mean (SD)** | 71.2 (11.5) | 70.8 (11.4) | 78.1 (10.1) | < 0.001 | 0.06 |
| Range | 35.8 – 100.3 | 35.8 – 100.3 | 51.7 – 93.8 |  |  |
| **Sex** |  |  |  | 0.623 | 0.34 |
| Female, n (%) | 606 (50.5) | 579 (50.7) | 27 (47.4) |  |  |
| Male, n (%) | 593 (49.5) | 563 (49.3) | 30 (52.6) |  |  |
| **Education (years), Mean (SD)** | 15.6 (2.4) | 15.6 (2.3) | 14.4 (2.6) | < 0.001 | < 0.001 |
| Range | 6 – 20 | 6 – 20 | 11 – 20 |  |  |
| **Device Type** |  |  |  | 0.964 | 0.72 |
| Desktop computer or laptop, n (%) | 751 (62.7) | 715 (62.7) | 36 (63.2) |  |  |
| Smartphone, n (%) | 266 (22.2) | 254 (22.3) | 12 (21.1) |  |  |
| Tablet, n (%) | 177 (14.8) | 168 (14.7) | 9 (15.8) |  |  |
| Other / not sure, n (%) | 4 (0.3) | 4 (0.3) | 0 (0.0) |  |  |
| Missing, n (%) | 1 (0.1) | 1 (0.1) | 0 (0.0) |  |  |
| **Session duration (min), Mean (SD)** | 16.4 (3.8) | 16.2 (3.6) | 19.3 (4.9) | < 0.001 | 0.77 |
| Range | 9.0 – 40.2 | 9.0 – 40.2 | 11.6 – 31.4 |  |  |
| **SLS Warm-Up duration (min), Mean (SD)** | 0.3 (0.3) | 0.3 (0.3) | 0.4 (0.3) | 0.010 | 0.007 |
| Range | 0.1 – 8.1 | 0.1 – 8.1 | 0.1 – 2.2 |  |  |
| **SLS Trials 1-5 duration (min), Mean (SD)** | 8.7 (2.1) | 8.7 (2.1) | 8.7 (2.5) | 0.828 | 0.74 |
| Range | 3.7 – 23.6 | 3.7 – 23.6 | 4.5 – 15.4 |  |  |
| **SLS Delay duration (min), Mean (SD)** | 1.4 (0.6) | 1.4 (0.6) | 1.5 (0.6) | 0.828 | 0.68 |
| Range | 0.5 – 10.0 | 0.5 – 10.0 | 0.6 – 3.7 |  |  |
| **Symbols Warm-Up duration (min), Mean (SD)** | 0.5 (0.3) | 0.4 (0.3) | 0.7 (0.5) | < 0.001 | 0.09 |
| Range | 0.1 – 3.6 | 0.1 – 3.6 | 0.2 – 3.1 |  |  |
| **Symbols Test duration (min), Mean (SD)** | 3.5 (1.1) | 3.5 (1.0) | 5.3 (1.9) | < 0.001 | < 0.001 |

| Range | 1.5 – 11.5 | 1.5 – 11.5 | 2.9 – 11.3 |
| --- | --- | --- | --- |

<sup>a</sup> Continuous variable p-values from linear model ANOVAs, categorical p-values from Pearson's Chi-Squared test.
<sup>b</sup> Adjusted p-values are adjusted for the effects of age, sex, and education using multivariable logistic regression models
*Note.* MTD = Mayo Test Drive, SD = standard deviation, SLS = Stricker Learning Span. See supplemental online materials for duration definitions. Table used with permission of Mayo Foundation for Medical Education and Research; all rights reserved.

**Table 3.**
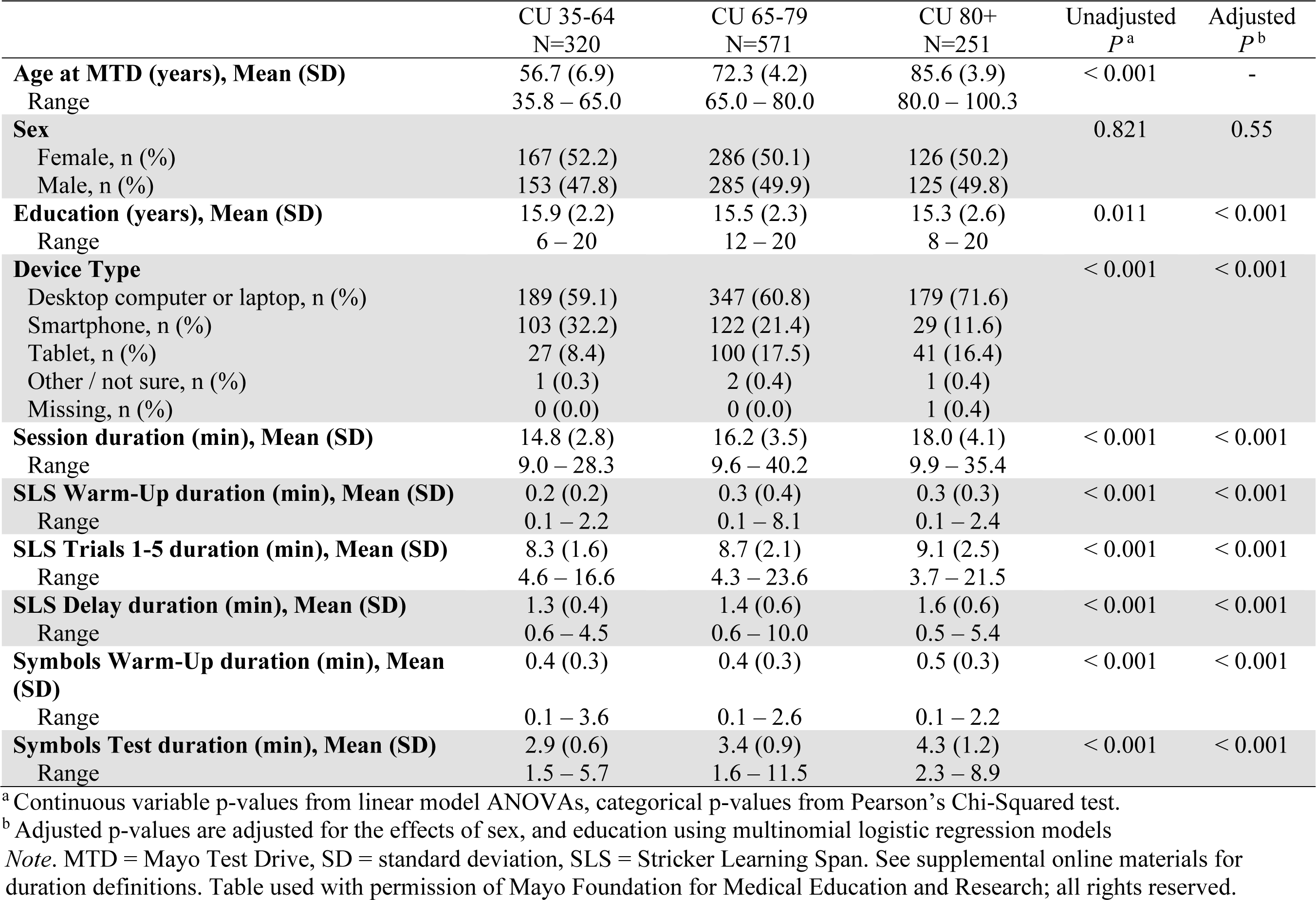
Session characteristics for Cognitively Unimpaired participants who completed a baseline MTD session by age subgroups.

**Table 4** reports several session characteristics relevant for understanding the test environment for remote sessions. Most participants completed testing at home (93%), 6% completed testing at work and <1% completed testing in a clinic or public space. Noise was endorsed in 4% of sessions; approximately half was noise that did not distract the participant. Subtests with longer durations had a higher percentage of participants reporting interference during that subtest (8.8% SLS learning trials, 5.7% Symbols, 1.8% SLS delay; see **Table 4** for response categories endorsed after a yes response about potential interference). All participants passed the SLS warm-up on the first (98%) or second (2%) attempt, suggesting participants were able to understand and follow instructions. While most also passed the Symbols warm-up on the first (92%) or second (6%) attempt, a few required a third attempt (0.5%) and one individual failed the warm-up and thus was not administered the full subtest (met discontinue rule).

**Table 4.** Location of testing and frequency of noise and subtest interference during initiated baseline sessions (N=1217).

| Variable | N (%) |
| --- | --- |
| **Context of session** |  |
| Location reported (N=1217) |  |
| *At home* | 1126 (92.5%) |
| *At work* | 71 (5.8%) |
| *In a clinic (medical or research center)* | 11 (0.9%)<sup>a</sup> |
| *In a public space (park, library)* | 9 (0.7%) |
| Noise in testing environment (N=1198) | 50 (4.2%) |
| *There was some noise in the background, but it did not distract me* | 27 (2.3%) |
| *There was some noise in the background and it was distracting* | 17 (1.4%) |
| *People were talking to me while I tried to take the test* | 2 (0.2%) |
| *People were talking in the background* | 4 (0.3%) |
| Validity/screen size concerns<sup>b</sup> | 12 (1.0%) |
| SLS practice “Warm-Up” pass rates (N=1217) |  |
| *SLS warm-up passed on first try (4/4)* | 1196 (98.3%) |
| *SLS warm-up passed on second try (3/4)* | 21 (1.7%) |
| *SLS warm-up passed on third try (2/4)* | 0 (0.0%) |
| *SLS warm-up failed (0 or 1 / 4)* | 0 (0.0%) |
| SYM practice “Warm-up” pass rates (N=1201) |  |
| *SYM warm-up passed on first try (4/4)* | 1123 (92.3%) |
| *SYM warm-up passed on second try (3/4)* | 71 (5.8%) |
| *SYM warm-up passed on third try (2/4)* | 6 (0.5%) |
| *SYM warm-up failed (0 or 1 / 4)* | 1 (0.1%) |
| **Interference endorsed during subtests** |  |
| SLS Trials 1-5 interference endorsed (N=1200) | 105 (8.8%) |
| *I am not comfortable using technology* | 2 (0.2%) |
| *I had technical problems* | 5 (0.4%) |
| *I was confused about the instructions* | 1 (0.1%) |
| *I was interrupted during this test* | 56 (4.6%) |
| *Sometimes my selection did not register* | 5 (0.4%) |
| *The words were hard for me to see* | 1 (0.1%) |
| *Other (there will be a comments box at the end of the session)* | 35 (2.9%) |
| SLS Delay interference endorsed (N=1198) | 22 (1.8%) |
| *I am not comfortable using technology* | 1 |
| *I had technical problems* | 1 |
| *I was confused about the instructions* | 0 |
| *I was interrupted during this test* | 2 |
| *Sometimes my selection did not register* | 4 |
| *The words were hard for me to see* | 1 |
| *Other (there will be a comments box at the end of the session)* | 13 |
| Symbols Test Interference endorsed (N=1198) | 69 (5.7%) |
| *I am not comfortable using technology* | 1 |
| *I had technical problems* | 5 |
| *I was confused about the instructions* | 3 |
| *I was interrupted during this test* | 15 |
| *Sometimes my selection did not register* | 13 |
| *The symbols were hard for me to see* | 1 |
| *Other (there will be a comments box at the end of the session)* | 30 |
| *Missing* | 1 |

<sup>a</sup> Seven participants elected to come into the clinic for a scheduled visit to do their MTD session. One of those individuals did not complete the session after they accidentally opened another window during SLS warm-up and then discontinued. We assume that the 4 additional individuals who endorsed their location as in clinic may work in a clinic or research center.
<sup>b</sup> Validity concerns are defined as present if there are no responses to items in the fourth position on any SLS learning trials (% responses in 4<sup>th</sup> position across all 5 trials = 0). This suggests that the 4<sup>th</sup> word displayed may not have been viewed. The response order for all warm-up trials was subsequently changed on 11/13/22 to help ensure the 4<sup>th</sup> word is always viewed by moving the correct response for all warm-up trials to the 4<sup>th</sup> position, making it impossible to pass the warm-up without selecting the response from the 4<sup>th</sup> position.
*Note.* SLS = Stricker Learning Span; SYM = Symbols Test. Table used with permission of Mayo Foundation for Medical Education and Research, all rights reserved.

Most participants who completed an MTD session used a personal computer (*n*=751, 62.7%). Many used a smartphone (n=266, 22.2%) or tablet (n = 177, 14.8%). Device use rates did not differ across cognitively impaired and unimpaired groups (**Table 2**, see **Supplemental Table 5** for MCI and dementia subgroups). However, there were differences in device use rates across age groups among CU participants (p < 0.001), with higher PC use and lower smartphone use in older age groups (**Figure 2**, **Table 3**).

**Figure 2.**
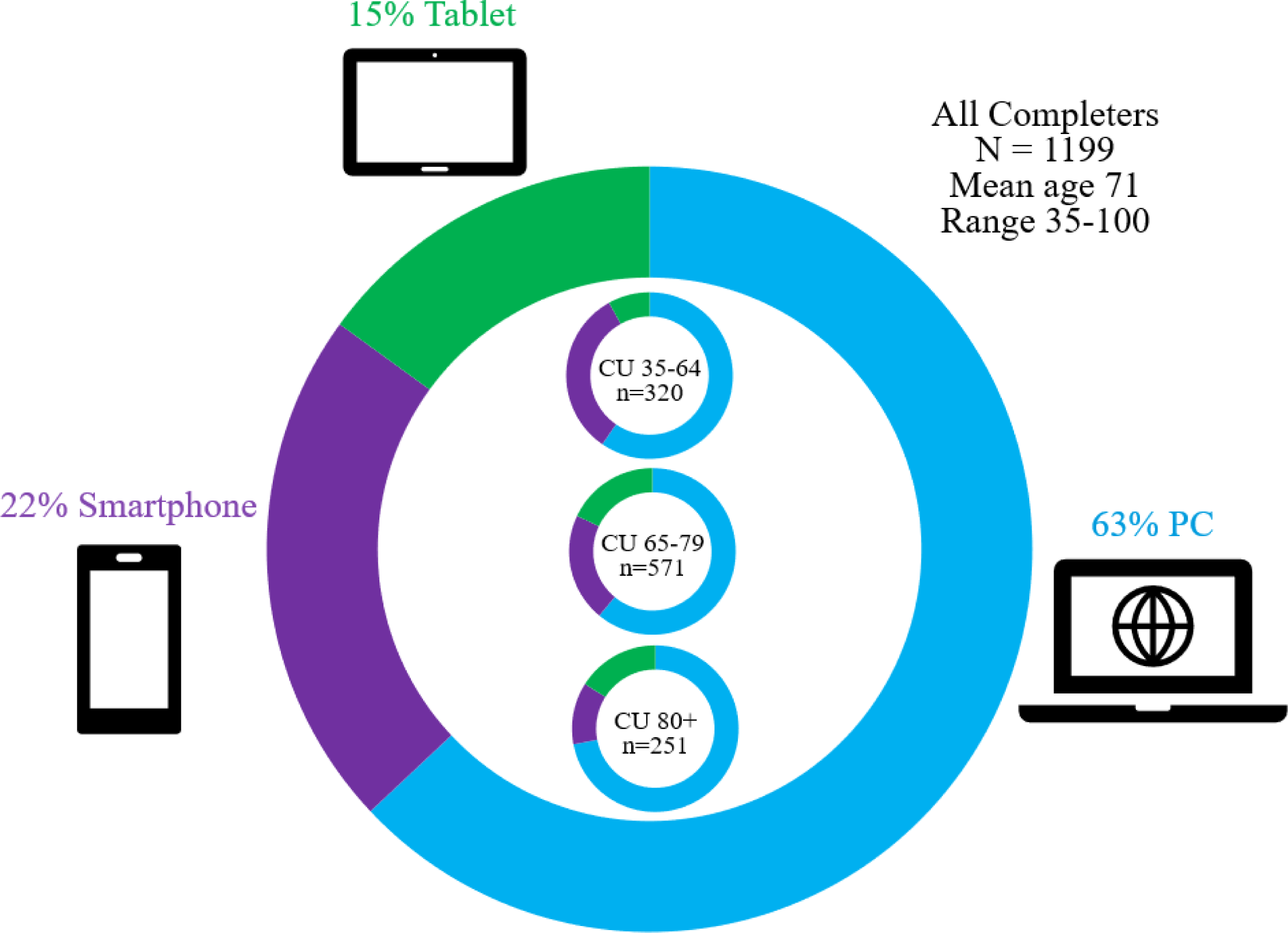
Device use for all participants who completed a session (large circle) and for Cognitively Unimpaired participants by age group (small circles). *Note*. Figure used with permission of Mayo Foundation for Medical Education and Research, all rights reserved.

Several session characteristics for the MTD follow-up session are presented in **Supplemental Table 6** and show a similar pattern of results.

### Qualitative Analysis of Participant Comments

Comments were entered voluntarily by 36.4% of participants (437/1199 participants completing a baseline session). Most comments were coded under one theme. Some longer comments contained multiple components that were coded into more than one theme, resulting in a total number of 481 comment components coded from 437 comments. See **Figure 3** for an overview of themes with examples, and **Table 5** for additional details and number of comments within theme subcategories.

**Figure 3.**
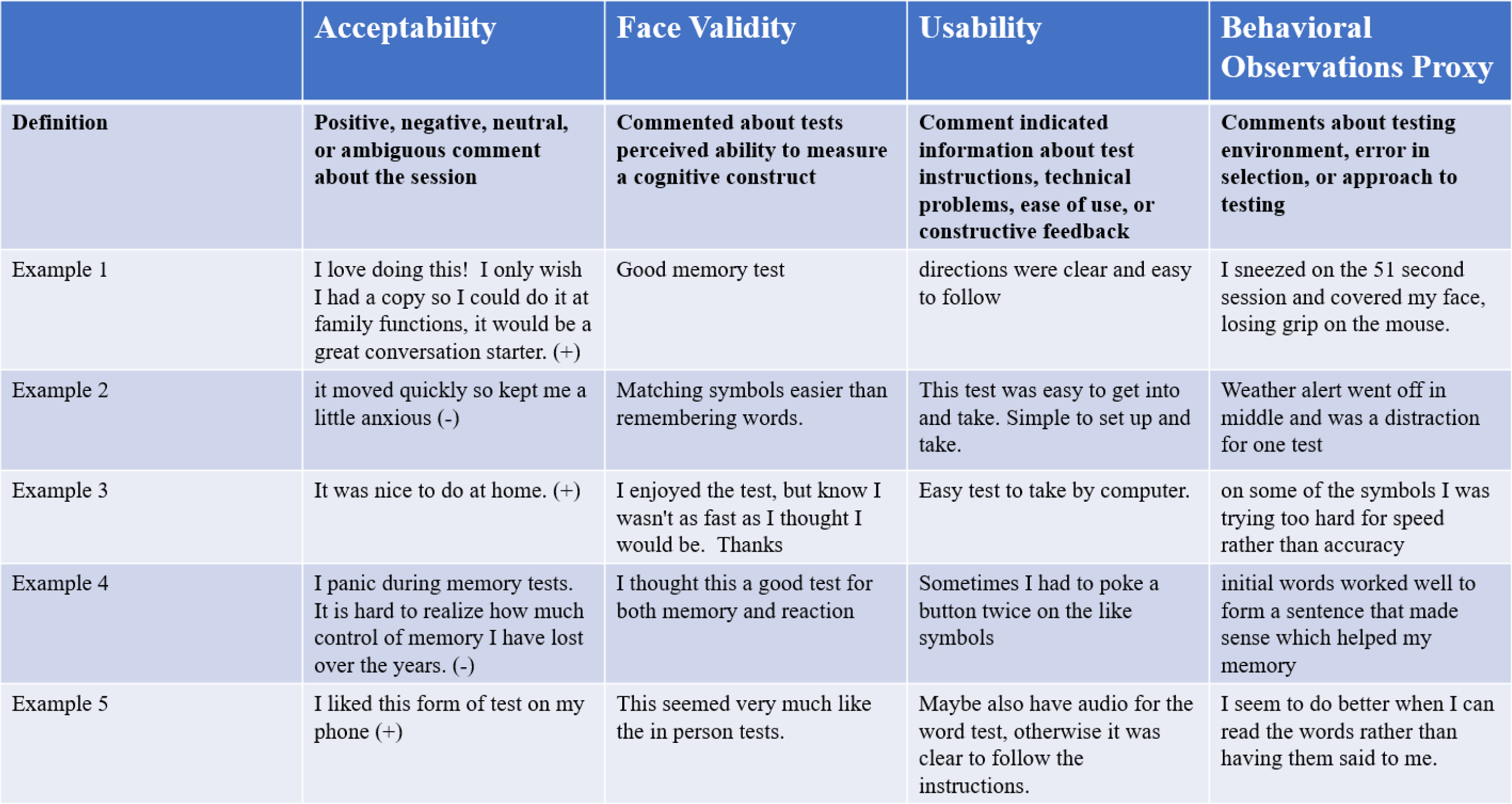
Comment rating categories overview and examples. *Note*. Figure used with permission of Mayo Foundation for Medical Education and Research, all rights reserved.

**Table 5.** Quantitative summary of qualitative review and groupings for optional comments provided during baseline session.

|  |  |
| --- | --- |
| **Completed Baseline Session, N** | 1199 |
| Total number of comments | 437 (36.4%) |
| *Number of comments coded into 1 theme* | 396 |
| *Number of comments coded into 2 themes* | 39 |
| *Number of comments coded into 3 themes* | 1 |
| *Number of comments coded into 4 themes* | 1 |
| Total number of comment elements coded into themes | 481 |
| *Comments coded as relating to behavioral observations proxy (theme 1)* | 86 (17.9%) |
| *Comments coded as relating to usability (theme 2)* | 67 (13.9%) |
| *Comments coded as relating to face validity (theme 3)* | 35 (7.3%) |
| *Comments coded as relating to acceptability (theme 4)* | 293 (60.1%) |
| **Comments coded as relating to Behavioral observations proxy (theme 1)** | 86 |
| Behavioral 1: | 61 |
| *Comment that provided useful information about the environment, context of the session, relevant internal or external distractions, etc. (e.g., there was a loud truck outside; my phone rang, usage due to health problems)* |  |
| Behavioral 2: | 7 |
| *Explained an incorrect selection (sometimes people endorsed interference when there was none; e.g., info that helps us understand their remote data better, participant stated they hit "yes" instead of "no" for interference.)* |  |
| Behavioral 3: | 4 |
| *Discussed their strategy / approach to the testing* |  |
| Behavioral 4: | 14 |
| *Comment that explained a response selection made, such as more specifics about response input used or how their choice of device/response input may have impacted performance* |  |
| **Comments coded as relating to usability (theme 2)** | 67 |
| Usability 1: | 5 |
| *Indication that instructions were easy to understand* |  |
| Usability 2: | 0 |
| *Indication that instructions were hard to understand* |  |
| Usability 3: | 1 |
| *Indication that they were slow to understand the task, were confused about instructions or expectations* |  |
| Usability 4: | 1 |
| *Easy to use comments* |  |
| Usability 5: | 20 |
| *Technical problems described* |  |
| Usability 6: | 3 |
| *Sensory or motor (health condition) impacts use of MTD* |  |
| Usability 7:<br>*Helpful constructive feedback* | 20 |
| Usability 8:<br>*Comments about length of testing* | 1 |
| **Comments coded as relating to face validity (theme 3)** | 35 |
| Face Validity 1:<br>*Indication that the test appeared to assess memory* | 25 |
| Face Validity 2:<br>*Complaint that the test was not assessing memory well* | 1 |
| Face Validity 3:<br>*Indication that Symbols test assesses speed of thinking, attention/working memory, or visual matching* | 7 |
| Face Validity 4:<br>*Requests for brain training exercises or other resources after completing the test, or indication that this test would be good for brain training exercise* | 3 |
| Face Validity 5:<br>*Other endorsement of face validity not falling into a category above* | 0 |
| **Comments coded as relating to acceptability (theme 4)** | 293 |
| **Comments coded as relating to acceptability (positive valence)** | 268 |
| Acceptability 1:<br>*Interesting* | 37 |
| Acceptability 2:<br>*Fun, enjoyed* | 63 |
| Acceptability 3:<br>*Good test (or similar)* | 21 |
| Acceptability 4:<br>*Reference to test being engaging or useful* | 1 |
| Acceptability 5:<br>*Hard, tough, challenging* | 66 |
| Acceptability 6:<br>*Thank you* | 47 |
| Acceptability 7:<br>*Positive comment about digital or remote format (nice to do at home; like doing on smartphone)* | 22 |
| Acceptability 8:<br>*Other positive response* | 17 |
| Acceptability 15:<br>*Comment about feedback provided being helpful* | 3 |
| Acceptability 17:<br>*Preference for digital self-administered test compared to in-person testing* | 2 |
| Acceptability 18:<br>*Preference for the new tests (MTD) or statement that they perceive the new tests as better compared to the “old tests” (cards, Cogstate)* | 24 |
| **Comments coded as relating to acceptability (negative valence)** | 12 |
| Acceptability 9:<br>*Comment suggesting test made them anxious* | 4 |
| Acceptability 11:<br>*Frustrating, discouraging* | 2 |
| Acceptability 12:<br>*Negative comment about digital or remote format (don't like doing it at home, etc.)* | 0 |
| Acceptability 13:<br>*Other negative response* | 3 |
| Acceptability 19:<br>*Preference for “old tests” (cards, Cogstate)* | 3 |
| **Comments coded as relating to acceptability with ambiguous significance** | 25 |
| Acceptability 14:<br>*Neutral response (or subjective judgement required and unclear how to rate, like with sarcasm)* | 10 |
| Acceptability 16:<br>*Requests for normative feedback; wondering how they did compared to others* | 2 |
| Acceptability 21:<br>*Ambiguous* | 9 |
| Acceptability 22:<br>*Other, comment not related to the test session, does not fit with any other rating options* | 4 |

*Note.* There are no acceptability 10 or 20 categories; those numbers were avoided to facilitate coding. Table used with permission of Mayo Foundation for Medical Education and Research, all rights reserved.

Most comments (60.1%) were related to acceptability; these were further categorized into perceived valence categories. A majority were positive (73% of acceptability comments, e.g., interesting, fun, challenging, enjoying digital or remote format, etc.), some were negative (12%; e.g., frustrating, feelings of anxiousness, etc.), and remaining comments were neutral (8.5%; e.g., requests for normative feedback, ambiguous comment, etc.).

About 18% of comments were categorized under the behavioral observations proxy theme. This theme represented comment categorizations relating to participants explaining relevant factors that may have influenced their performance during testing, their testing approach, and explanations regarding response selection.

Approximately 14% of comment categorization represented usability. Some participants spontaneously stated instructions were easy to understand (*n*=5). Several described technical problems (n=20; 4.2% of all comment categorizations or 1.7% of all baseline sessions completed). Several provided helpful constructive feedback (*n*=20), some of which were used to make platform enhancements.

Finally, several comments related to face validity (*n*=35, 7.3% of comments) and predominantly supported the face validity of the subtests.

## Discussion

This study supports the usability, feasibility, and acceptability of the remotely administered MTD screening battery. We demonstrate high usability of MTD, with 98.5% completion rates and no significant differences in those with and without cognitive impairment or across age groups. Follow-up adherence is also high (89%). MTD self-administered sessions are completed efficiently (mean 16 minutes) using multiple device types. Voluntary participant comments provide additional support for MTD acceptability and face validity.

Objective usability results support our primary aim, with 98.5% of participants who initiated a session completing a session. This exceeded our hypothesized goal of 90% completion rates. Our high completion rates are similar to those reported for a younger research cohort using the Cogstate Brief Battery (CBB); Perrin et al. [6] reported 98% completion rates in a sample of participants aged 40-65 (mean=56, SD 7). Our completion rates are also comparable to a self-administered computerized screening measure administered unsupervised in clinic waiting rooms; specifically, the Cleveland Clinic Cognitive Battery (C3B) showed >92% completion rates in a primary care clinic [33]. Ashford et al. [34] found that 80.8% of Brain Health Registry (BHR) participants who started the Paired Associates Learning Task from the Cambridge Neuropsychological Test Automated Battery® (CANTAB PAL) completed the task in a sample with a broad age range (18-99, mean=66, SD=11). The most common reasons for starting but not completing the PAL were technical difficulties and lack of device support (e.g., test was not able to be completed in the BHR on smartphones or tablets at that time).

MTD completion rates were comparable across cognitively unimpaired and cognitively impaired groups, supporting the high usability of the platform. This finding is notable because completion rates for some examiner-administered computerized tasks completed in clinic have shown usability limitations in cognitively impaired participants [3]. Results also showed generally comparable completion rates across age groups, ranging from 97.3% in individuals aged 80+ to 99.7% in individuals aged 34-64. Several MTD design characteristics likely helped maintain these high completion rates in older adults, including multi-device compatibility and no use of audio. Hischa et al. [13] highlighted that use of headphones for a tablet-based, self-administered battery of working memory tests frequently interfered with hearing aids, and this may have resulted in reduced completion rates across age groups (e.g., 32% of oldest-old participants completed the tests vs. 97% of young adults). Requiring audio could similarly lead to unexpected difficulties in individuals with hearing aids for measures developed for primary care settings that use headphones to facilitate administration in waiting rooms [33]. While there are some benefits to presenting auditory instructions, we chose to avoid use of audio for the MTD screening battery to enhance usability because hearing loss is common in older adults. Marinelli et al. [15] reported that among 1200 CU individuals in the Mayo Clinic Study of Aging (MCSA; mean age 76, SD=9) who volunteered to undergo formal behavioral audiometric evaluation by an audiologist, only 36% had normal hearing and other participants had mild (32%), moderate (30%) or severe/profound (1%) hearing loss. Further, Gorman et al. [35] showed that rates of moderate hearing loss increase substantially in individuals aged 80+ (38% prevalence) relative to younger age groups (16% in 70-79 year-olds; 6% in 60-69 year-olds and 2% or below in those <60 years). Requiring audio also increases technological complexity and may be a barrier to participation if audio is not set up correctly. The differing quality of output of different devices and peripherals may also lead to added variability. A tradeoff of the decision to avoid use of audio to increase accessibility for people with hearing impairment is that MTD therefore requires vision. Because participants can choose their preferred device to take MTD, individuals with vision loss responsive to accommodations may elect to use a desktop computer or tablet that allows for a larger screen size and text size than a smartphone. In addition, we considered design needs for older adults during test development and prioritized large font (e.g., particularly for memory test items) and simple high contrast visual displays.

Although results show that the MTD platform is easy to use, our data illustrate a common pattern observed in computerized and remote cognitive assessment studies wherein individuals with cognitive impairment and those aged 80+ years are less likely to participate in technology-based studies, particularly on their own without assistance. Similar to survey data showing that only 39% of participants with dementia reported willingness to engage in remote testing [4], our results showed that the percentage of participants willing to attempt participation in this uncompensated, remote cognitive assessment ancillary study was lower for those with cognitive impairment (33%) compared to CU individuals (65%). This is consistent with other studies examining observed rates of engagement with voluntary, unsupervised, self-administered neuropsychological measures. For example, Weiner et al. showed that a self-reported diagnosed memory problem was associated with decreased likelihood of completing self-administered neuropsychological tests in the BHR [36]. Even among individuals registering for an online platform such as the BHR, many do not complete available self-administered neuropsychological tests. In a recent update, Weiner et al., reported that 56% of BHR participants completed at least one neuropsychological test and 22% completed at least two neuropsychological tests at baseline [37]. Ashford et al. [34] reported that 50.6% of BHR participants who were provided an opportunity to complete the CANTAB PAL did not attempt it. In our prior work with the CBB in the MCSA, we administered the CBB during each in person study visit and allowed participants the option to complete an interim session(s) in clinic or remotely. Using that study design, only 18% of MCI participants ever completed an at home Cogstate session and 17% of all MCSA participants never completed a CBB session (in clinic or at home; unpublished data).

Once enrolled, we found that initial adherence/retention rates for the 7-8 month MTD follow-up session were 89%, with 99% completion rates for initiated follow-up sessions. This suggests individuals are willing to re-engage with the MTD platform, supporting feasibility of longitudinal monitoring. These retention rates are promising given that retention is often lower than desired in remote cognitive assessment studies. For example, in an ADNI pilot study, after initially completing CBB in clinic, 79.4% of CU and MCI participants completed the first follow-up (within 2 weeks) and 37.1% completed a 6-month follow-up. In the Alzheimer Prevention Trials web-study, Walter et al. [11] similarly reported that a relatively small number of individuals completed follow-up remote computerized testing, with 29.5% completing a second CBB session, and 23% completing a third CBB session. Ashford et al. [38] recently demonstrated that the presence and number of self-reported medical conditions impacts longitudinal completion of questionnaires and the CBB in the BHR. Overall, they found that 75% of individuals 55+ completed a questionnaire at least twice, and 45% completed the CBB at least twice. The number of self-reported medical conditions was negatively associated with likelihood of completing at least two cognitive assessments but was not associated with likelihood of longitudinal questionnaire completion. Those specifically self-reporting a history of Alzheimer’s disease and related disorders were less likely to complete either the questionnaire or the CBB twice.

Results from registry studies suggest that platform flexibility in terms of device compatibility may be a critical factor underlying both willingness to participate and usability [11, 34]. For example, Walter et al. [11] reported that 97% of participants consenting to the APT web-study completed a self-report questionnaire of cognitive symptoms (the Cognitive Function Index, CFI) but that only 65% completed initial remote self-administered computerized testing (CBB). The difference in rates of self-report versus computerized testing completion in that study was described as due in part to technical challenges and lack of compatibility of the CBB with smartphones. Our group previously administered the CBB in the MCSA and study coordinators received many requests for a smartphone compatible option. Despite this feedback, a majority (63%) of participants in our study used a personal computer to complete MTD, and 37% used a mobile device (22% smartphone, 15% tablet). Smartphone use for completing MTD was higher in younger relative to older age groups; conversely, personal computer use increased with age. These results suggest that the target demographics of a study or clinic may influence device preference. Device flexibility may be important to ensure ability of users to complete assessments [11, 34].

“Bring your own device” (BYOD) approaches may engage the most users, even though there can be some minor psychometric disadvantages with use of multiple device types [27, 39]. BYOD often implies choosing a device type within a given device class, such as choosing either android or iOS devices in a smartphone-only study. The ability to choose the device participants are most comfortable with, which we suggest represents a “Choose Your Preferred Device” (CYPD) approach, is likely particularly important for individuals over 80 and for those with cognitive impairment. These groups are likely to be more hesitant to participate in self-administered remote cognitive assessment studies based on our results; therefore, the ability to use a familiar device may increase participation rates. Many studies directly supply devices to participants. Although this helps overcome any barriers regarding device ownership and usage, this may inadvertently introduce differential impact of ability or willingness to adapt to an unfamiliar device and add burden to study personnel or participants for device delivery. A CYPD approach ensures participants can complete cognitive testing on a device they are most comfortable with. This approach also best aligns with real-world settings such as clinical practices making use of remote assessment technology or pragmatic clinical trials. Another potential explanation for the higher-than-expected use of personal computers in the current study is that participants may be most likely to use the device they use to check email, since email was the primary method of providing links to the test. An option to allow participants without an available device to come to the clinic to complete testing may be one strategy to mitigate potential bias; this is a strategy employed in the current study, though very few participants made use of this option when it was available.

Several additional factors beyond device flexibility inform feasibility for implementing unsupervised remote digital cognitive assessments in research and clinical settings, including session duration and frequency of interruptions during test sessions. Our prior MTD pilot study showed that an MTD session was typically completed in 15 minutes (median) in older adult females, whose mean age was 5 years younger than the current sample. The current results are similar, with mean session duration of 16 minutes. To aid group comparisons, we reported means instead of medians in the current study, though means will show slightly longer durations than medians. Mean subtest durations were 8.7 minutes for SLS learning trials, 3.5 minutes for Symbols trials, and 1.4 minutes for SLS delay. There were significantly longer durations for the total session and each subtest in older age groups. The cognitively impaired group showed slower completion times for the total session and for Symbols relative to the CU group. However, there was no difference in duration across CU and CI groups for SLS learning trials, likely due to the adaptive nature of the SLS that often results in fewer items presented to CI participants. Most participants reported completing sessions at home (93%). Noise was infrequently reported (4% of session). Participants endorsed interference during subtests in 2-9% of sessions; the frequency of this varied by subtest duration, with longer subtests having a higher likelihood of interference. Most participants were able to correctly answer the practice item on the first attempt, suggesting they were able to understand task instructions.

Qualitative analysis of voluntary comments left at the end of MTD sessions support acceptability, with a majority of positive comments. Qualitative analyses also provided evidence for face validity of subtests. As described by Soobiah et al. (2019), face validity is important for implementation of self-administered measures because participants may be more likely to complete the measure if they perceive it to have value/face validity. Comment analyses showed that 25 participants spontaneously made a comment indicating that the test appeared to assess memory. Many participants provided comments helpful for individual level interpretation of their test performance (behavioral observations proxy), such as describing a distraction that occurred or stating they accidentally endorsed an interruption when in fact there was none. A minority of participant comments described technical problems (1.7% of all baseline sessions). Many participants also provided helpful constructive feedback, including some that led to platform enhancements. Thus, in addition to an avenue for learning important information about the context of sessions directly from participants in these remote, unsupervised sessions, free-text comments also provide a method of ongoing participant feedback. While cognitive interviewing approaches during the test development phase can accomplish similar goals [14], free-text comments provide a low burden opportunity for all individuals taking the tests across all use cases to provide this ongoing, valuable input.

The current study has some limitations to consider. First, the sample is predominantly White, Non-Hispanic. Future work is needed to determine if results generalize to a more sociodemographically diverse sample. Our current R01 includes a focus on increasing representation of African American participants in this work, and we are also working on a Spanish adaptation of MTD [40]. Second, examining usability, feasibility and acceptability in research participants asked to complete a session of MTD for a voluntary research study without compensation may not generalize to broader contexts such as those seeking medical attention surrounding cognitive concerns. For example, while the participation rates observed among cognitively impaired participants in this study may raise some concerns about the acceptability of any digital cognitive measure being regularly implemented in clinical settings, participation rates in this study reflect willingness to voluntarily engage in this ancillary research study and may not reflect willingness to engage with a remote cognitive assessment when it is requested by a clinical provider. Clinically referred remote cognitive assessments may have higher participation rates due to higher motivation to complete a remote, cognitive test (e.g., in settings with low base rates of cognitive impairment such as primary care clinics). Conversely, lower clinical participation and completion rates could be seen because of higher base rates of cognitive impairment in some settings (e.g., dementia clinic). Future studies are needed to examine usability, feasibility, and acceptability findings in clinical care settings. Based on cognitive interview data, Young et al. [14] found that clinicians, administrators, and healthy older adults preferred remote, pre-visit cognitive screening because it saved time relative to cognitive screening measures that require person administration and scoring. However, this was based on theoretical feedback and opinions and not on objective data inviting patients to complete pre-visit remote cognitive assessments. It is also possible that participants with cognitive impairment may be less likely to check their email, which was the primary method of communication in this remote study.

Overall, the MTD platform demonstrates high usability in this research sample that includes representation of a broad age range, with good representation of individuals over 80 and inclusion of some individuals with cognitive impairment. Though likelihood of participation in remote cognitive assessment studies appears to vary by cognitive status and age, further research is needed to understand generalizability in clinical settings. The current results add to our prior work establishing the concurrent and criterion validity of MTD [17–19, 29] and show support for the feasibility, acceptability, and face validity of MTD platform as well as the Stricker Learning Span and Symbols subtests.

## Funding and Acknowledgments

This research was supported by the National Institute on Aging of the National Institutes of Health (R01 AG081955, R21 AG073967, P30 AG062677, U01 AG006786, RF1 AG069052), the Kevin Merszei Career Development Award in Neurodegenerative Diseases Research IHO Janet Vittone, MD, the Rochester Epidemiology Project (NIH R01 AG034676), the GHR Foundation, and the Mayo Foundation for Education and Research. The content is solely the responsibility of the authors and does not necessarily represent the official views of the National Institutes of Health or other funding sources. The authors wish to thank the participants and staff at the Mayo Clinic Study of Aging and Mayo Alzheimer’s Disease Research Center.

## Data Availability

The data supporting the findings of this study are available upon reasonable request to the corresponding author and with approval from Mayo Clinic Study of Aging investigators.

