## Supplemental Online Materials for "Usability of the Mayo Test Drive remote self-administered web-based cognitive screening battery in adults ages 35 to 100 with and without cognitive impairment"

### Supplemental Online Resources

Jay S. Patel [1], Teresa J. Christianson [3], Logan T. Monahan, [1] Ryan D. Frank [3], Winnie Z. Fan [3], John L. Stricker [2], Walter K. Kremers [3], Aimee J. Karstens [1], Mary M. Machulda [1], Julie A. Fields [1], Jason Hassenstab [4], Clifford R. Jack, Jr. [5], Hugo Botha [6], Jonathan Graff-Radford [6], Ronald C. Petersen [6], Nikki H. Stricker [1]

[1] Division of Neurocognitive Disorders, Department of Psychiatry and Psychology, Mayo Clinic, Rochester, Minnesota, USA
[2] Department of Information Technology, Mayo Clinic, Rochester, Minnesota, USA
[3] Division of Biomedical Statistics and Informatics, Department of Quantitative Health Sciences, Mayo Clinic, Rochester, Minnesota, USA
[4]Department of Neurology and Psychological & Brain Sciences, Washington University in St. Louis, St. Louis, Missouri, USA
[5] Department of Radiology, Mayo Clinic, Rochester, Minnesota, USA
[6] Department of Neurology, Mayo Clinic, Rochester, Minnesota, USA

Copyright 2024 Mayo Foundation for Medical Education and Research, all rights reserved.

**Supplemental Table 1.** Participation and baseline session completion rates for individuals invited to participate in the Mayo Test Drive between 5/25/21 and 10/4/22.

| | Total Sample N=1950 | Cognitively Unimpaired N=1769 | Cognitively Impaired N=181[a] | Unadjusted *P*[b] | Adjusted *P*[c] |
|---|---|---|---|---|---|
| **Participation Rates** | 1217 (62.4%) | 1157 (65.4%) | 60 (33.1%) | <.001 | <.001 |
| **Session Completion** | 1199 (98.5%) | 1142 (98.7%) | 57 (95.0%) | .049 | .23 |
| **Reason for non-participation** | | | | | |
| No response | 503 (67.0%) | 434 (69.2%) | 69 (55.6%) | - | - |
| Declined | 233 (31.1%) | 183 (29.2%) | 50 (40.4%) | - | - |
| Death within 6 weeks of invite | 2 (0.3%) | 1 (0.2%) | 1 (0.8%) | - | - |
| Not willing to attempt or unable | 4 (0.5%) | 4 (0.6%) | 0 (0.0%) | - | - |
| No access to tech. | 9 (1.2%) | 5 (0.8%) | 4 (3.2%) | - | - |

[a] There were 35 participants with a dementia diagnosis invited to participate, 9 who initiated a session and 8 who completed. The remaining cognitively impaired participants had an MCI diagnosis.
[b] Unadjusted p-values from logistic regression
[c] Adjusted for the effects of age, sex, and education using logistic regression
*Note*. Table used with permission of Mayo Foundation for Medical Education and Research; all rights reserved.

**Supplemental Table 2.** Participation and baseline session completion rates for cognitively unimpaired individuals by age group.

| | CU 34-64 N=466 | CU 65-79 N=823 | CU 80+ N=480 | Unadjusted *P* [a] | Adjusted *P* [b] |
|---|---|---|---|---|---|
| **Participation Rates** | 321 (68.9%) | 578 (70.2%) | 258 (53.8%) | <.001 | <.001 |
| **Session Completion** | 320 (99.7%) | 571 (98.8%) | 251 (97.3%) | 0.11 | .098 |
| **Reason for non-participation** | | | | | |
| No response | 128 (87.7%) | 184 (73.0%) | 122 (53.3%) | - | - |
| Declined | 17 (11.6%) | 67 (26.6%) | 99 (43.1%) | - | - |
| Death within 6 weeks of invite | 1 (0.7%) | 0 (0%) | 0 (0%) | - | - |
| Not willing to attempt or unable | 0 (0%) | 1 (0.4%) | 3 (1.3%) | - | - |
| No access to technology | 0 (0%) | 0 (0%) | 5 (2.2%) | - | - |

[a] Unadjusted p-values from logistic regression

[b] Adjusted for the effects of sex and education using multinomial logistic regression

*Note*. Table used with permission of Mayo Foundation for Medical Education and Research; all rights reserved.

**Supplemental Table 3.** Participation and baseline session completion rates for cognitively impaired individuals by diagnostic group.

| | MCI N=146 | Dementia N=35 | Unadjusted *P* [a] | Adjusted *P* [b] |
|---|---|---|---|---|
| **Participation Rates** | 51 (34.9%) | 9 (25.7%) | 0.301 | .298 |
| **Session Completion** | 49 (96.1%) | 8 (88.9%) | 0.383 | .362 |
| **Reason for non-participation** | | | | |
| No response | 55 (56.7%) | 14 (51.9%) | - | - |
| Declined | 37 (38.2%) | 13 (48.1%) | - | - |
| Death within 6 weeks of invite | 1 (1.0%) | 0 (0%) | - | - |
| Not willing to attempt or unable | 0 (0%) | 0 (0%) | - | - |
| No access to technology | 4 (4.1%) | 0 (0%) | - | - |

[a] Unadjusted p-values from logistic regression

[b] Adjusted for the effects of age, sex, and education using logistic regression

*Note*. Table used with permission of Mayo Foundation for Medical Education and Research; all rights reserved.

**Supplemental Table 4.** Retention and follow-up completion rates for individuals invited to do a 7.5-month follow-up session (participants through 10/4/22)

| | | Cognitively Unimpaired | | | | | Cognitively Impaired | | | |
|---|---|---|---|---|---|---|---|---|---|---|
| | Total | Total | 30-64 | 65-79 | 80+ | *P* [a] | Total | MCI | Dementia | *P* [b] |
| Number follow-up emails | 583 | 558 | 141 | 261 | 156 | | 25 | 24 | 1 | |
| MTD Initiated | 519 (89.0%) | 503 (90.1%) | 119 (84.4%) | 246 (94.3%) | 138 (88.5%) | 0.006 | 16 (64.0%) | 15 (62.5%) | 1 (100%) | 0.901 |
| Session Completed | 514 (99.0%) | 500 (99.4%) | 117 (98.3%) | 245 (99.6%) | 138 (100%) | 0.649 | 14 (87.5%) | 13 (86.7%) | 1 (100%) | 0.866 |

[a] *P* value is adjusted for sex, and education using multinomial logistic regression models.
[b] *P* value is adjusted for age, sex, and education using logistic regression models.
*Note*. MTD = Mayo Test Drive. Table used with permission of Mayo Foundation for Medical Education and Research; all rights reserved.

**Supplemental Table 5.** Session characteristics for Cognitively Impaired participants who completed a baseline MTD session by diagnostic subgroups.

| | MCI N=49 | Dementia N=8 | Unadjusted *P* [a] | Adjusted *P* [b] |
|---|---|---|---|---|
| **Age at MTD (years), Mean (SD)** | 78.7 (10.2) | 74.6 (9.1) | .29 | .14 |
| Range | 51.7 – 93.8 | 64.1 – 87.2 | | |
| **Sex** | | | .87 | .80 |
| Female, n (%) | 23 (46.9) | 5 (50.0) | | |
| Male, n (%) | 26 (53.1) | 5 (50.0) | | |
| **Education (years), Mean (SD)** | 14.1 (2.4) | 16.3 (3.2) | .30 | .02 |
| Range | 11.0 – 20.0 | 12.0 – 20.0 | | |
| **Device Type** | | | .66 | .72 |
| Desktop computer or laptop, n (%) | 31 (63.3) | 5 (62.5) | | |
| Smartphone, n (%) | 11 (22.4) | 1 (12.5) | | |
| Tablet, n (%) | 7 (14.3) | 2 (25.0) | | |
| Other / not sure, n (%) | 0 (0.0) | 0 (0.0) | | |
| Missing, n (%) | 0 | 0 | | |
| **Session duration (min) , Mean (SD)** | 19.1 (4.9) | 20.0 (5.4) | .65 | .99 |
| Range | 11.6 – 31.0 | 16.0 – 31.4 | | |
| **SLS Warm-Up duration (min) , Mean (SD)** | 0.4 (0.2) | 0.5 (0.7) | .081 | .052 |
| Range | 0.1 – 1.0 | 0.2 – 2.2 | | |
| **SLS Trials 1-5 duration (min) , Mean (SD)** | 8.8 (2.5) | 8.4 (2.9) | .74 | .48 |
| Range | 4.5 – 15.4 | 4.7 – 13.6 | | |
| **SLS Delay duration (min) , Mean (SD)** | 1.5 (0.6) | 1.4 (0.4) | .51 | .28 |
| Range | 0.6 – 3.7 | 1.0 – 1.9 | | |
| **Symbols Warm-Up duration (min) , Mean (SD)** | 0.7 (0.5) | 0.9 (0.7) | .34 | .79 |
| Range | 0.2 – 3.1 | 0.2 – 2.3 | | |
| **Symbols Test duration (min) , Mean (SD)** | 5.1 (1.7) | 6.2 (2.4) | .11 | .28 |
| Range | 2.9 – 11.3 | 3.25 – 9.5 | | |

[a] Continuous variable *P* values from linear model ANOVAs, categorical p-values from Pearson's Chi-Squared test.
[b] *P* values adjusted for the effects of age, sex, and education using multivariable logistic regression models.
*Note*. MTD = Mayo Test Drive, SD = standard deviation, SLS = Stricker Learning Span. Table used with permission of Mayo Foundation for Medical Education and Research; all rights reserved.

**Supplement Table 6.** Location of testing and frequency of noise and subtest interference during initiated follow-up sessions, N=518.

| **Variable** | N (%) |
|---|---|
| **Context of session** | |
| Location reported (N=518) | |
| *At home* | 486 (93.8%) |
| *At work* | 28 (5.4%) |
| *In a clinic (medical or research center)* | 3 (0.6%) [a] |
| *In a public space (park, library)* | 1 (0.2%) |
| Noise in testing environment (N=514) | 16 (3.1%) |
| *There was some noise in the background, but it did not distract me* | 5 (1.0%) |
| *There was some noise in the background and it was distracting* | 5 (1.0%) |
| *People were talking to me while I tried to take the test* | 0 (0%) |
| *People were talking in the background* | 6 (1.2%) |
| Screen size concerns (SLS 1-5 correct with 0% responses in 4$^{th}$ position) | 8 (1.5%) |
| Practice "Warm-Up" pass rates | |
| *SLS warm-up passed on first try (4/4)* | 508 (98.1%) |
| *SLS warm-up passed on second try (3/4)* | 7 (1.4%) |
| *SLS warm-up passed on third try (2/4)* | 0 (0.0%) |
| *SLS warm-up failed (0 or 1 / 4)* | 0 (0.0%) |
| *SYM warm-up passed on first try (4/4)* | 482 (93.1%) |
| *SYM warm-up passed on second try (3/4)* | 25 (4.8%) |
| *SYM warm-up passed on third try (2/4)* | 5 (1.0%) |
| *SYM warm-up failed (0 or 1 / 4)* | 0 (0.0%) |
| **Interference endorsed during subtests** | |
| SLS Trials 1-5 interference endorsed (N=512) | 37 (7.1%) |
| *I am not comfortable using technology* | 2 (0.4%) |
| *I had technical problems* | 0 (0%) |
| *I was confused about the instructions* | 0 (0%) |

| | |
|---|---|
| *I was interrupted during this test* | 16 (3.1%) |
| *Sometimes my selection did not register* | 3 (0.6%) |
| *The words were hard for me to see* | 1 (0.2%) |
| *Other (there will be a comments box at the end of the session)* | 15 (2.9%) |
| SLS Delay interference endorsed (N=512) | 10 (1.9%) |
| *I am not comfortable using technology* | 0 (0%) |
| *I had technical problems* | 1 (0.2%) |
| *I was confused about the instructions* | 0 (0%) |
| *I was interrupted during this test* | 1 (0.2%) |
| *Sometimes my selection did not register* | 0 (0%) |
| *The words were hard for me to see* | 0 (0%) |
| *Other (there will be a comments box at the end of the session)* | 8 (1.6%) |
| Symbols Test Interference endorsed (N=512) | 39 (7.5%) |
| *I am not comfortable using technology* | 0 (0%) |
| *I had technical problems* | 4 (0.8%) |
| *I was confused about the instructions* | 0 (0%) |
| *I was interrupted during this test* | 8 (1.6%) |
| *Sometimes my selection did not register* | 6 (1.2%) |
| *The symbols were hard for me to see* | 3 (0.6%) |
| *Other (there will be a comments box at the end of the session)* | 18 (3.5%) |

[a] One individual elected to come into the clinic for a scheduled visit to do their follow-up MTD session. We assume that the 2 additional individuals who endorsed their location as in clinic may work in a clinic or research center.

*Note*. SLS = Stricker Learning Span; SYM = Symbols Test. Table used with permission of Mayo Foundation for Medical Education and Research; all rights reserved.

*Supplemental Methods.* Detailed definitions of duration variables are provided below.

Session duration = total time (minutes) from when a participant hits the “ready” button on the welcome screen to the presentation of the final screen that alerts participants that all tests are completed and invites them to provide comments (time spent writing comments was not included since that is optional).

Stricker Learning Span Warm-Up duration = total time (minutes) from when the “Memory Warm-Up” screen is presented to the presentation of the screen that alerts participants that the memory warm-up is completed (or that the memory test is completed if the warm-up is failed and the memory test is discontinued).

Stricker Learning Span Trials 1-5 duration = total time (minutes) from when the “What To Expect” screen is presented (the start of the instructions for Trials 1-5) to the presentation of the “Memory Test Complete” screen after trial 5.

Stricker Learning Span Delay duration = total time (minutes) from when the “Memory Check” screen is presented to the presentation of the “Memory Check Complete” screen after all delay items are tested.

Symbols Warm-Up duration = total time (minutes) from when the “Symbols Warm-Up” screen is presented to the presentation of the screen that alerts participants that the Symbols warm-up is completed (or that the Symbols test is completed if the warm-up is failed and the test is discontinued).

Symbols Test duration = total time (minutes) from when the “Symbols round 1 of 4” screen is presented (that provides a reminder of the instruction screen as also displayed for the Warm-Up) to the presentation of the “Symbols Test Complete” screen after trial 4.

*Note*. Session duration was screened for sessions > 30 minutes or occasionally based on interference reported and qualitative session review. When item level data was reviewed and showed that a screen was open for an abnormally long time, it was assumed that this represented a time when the participant was not taking the test and the time on that screen only was subtracted from the total duration for the purpose of this manuscript (14 sessions edited for the dates included in this manuscript). More than half (8/14) of these long pauses occurred on the “Welcome” screen or on the “Session Instructions” screen that includes instructions ensuring participants are in a quiet area without distractions for 15-20 minutes. For example, one participant was on the “Session Instructions” screen for 30 minutes. We assume these long pauses reflected that participants were following these instructions before resuming the session. Two sessions had long pauses during the instructions for SLS Trials 1-5. Four sessions had a long pause mid-task (e.g., instruction screen for one of the SLS trials). Based on this data, in future work we plan to use the time when a session is initiated (once a location is selected) as the session start time since this will address most of these long pauses and will provide a better representation of session duration in these instances.
